# Evaluating splatter and settled aerosol during orthodontic debonding: implications for the COVID-19 pandemic

**DOI:** 10.1101/2020.08.19.20178319

**Authors:** Hayley Llandro, James R Allison, Charlotte C Currie, David C Edwards, Charlotte Bowes, Justin Durham, Nicholas Jakubovics, Nadia Rostami, Richard Holliday

## Abstract

**Introduction:** Dental procedures produce splatter and aerosol which have potential to spread pathogens such as SARS-CoV-2. Mixed evidence exists on the aerosol generating potential of orthodontic procedures. The aim of this study was to evaluate splatter and/or settled aerosol contamination during orthodontic debonding.

**Material and Methods:** Fluorescein dye was introduced into the oral cavity of a mannequin. Orthodontic debonding was undertaken with surrounding samples collected. Composite bonding cement was removed using a speed-increasing handpiece with dental suction. A positive control condition included a water-cooled, high-speed air-turbine crown preparation. Samples were analysed using digital image analysis and spectrofluorometric analysis.

**Results:** Contamination across the 8-metre experimental rig was 3% of the positive control on spectrofluorometric analysis and 0% on image analysis. Contamination of the operator, assistant, and mannequin, was 8%, 25%, and 28% of the positive control, respectively.

**Discussion:** Splatter and settled aerosol from orthodontic debonding is distributed mainly within the immediate locality of the mannequin. Widespread contamination was not observed.

**Conclusions:** Orthodontic debonding is unlikely to produce widespread contamination via splatter and settled aerosol, but localised contamination is likely. This highlights the importance of personal protective equipment for the operator, assistant, and patient. Further work is required to examine suspended aerosol.

**Three ‘In brief’ points:**

- Orthodontic debonding, including removal of composite using a slow speed handpiece with dental suction, appears to pose little risk of widespread distribution of settled contamination.
- Splatter and settled aerosol was produced during the debonding procedure, however this was mainly localised to the patient, operator and assistant.
- Further work is required to examine aerosol which remains suspended in the air.

## Introduction

The delivery of orthodontics has changed rapidly over the last several months as a result of the coronavirus disease 2019 (COVID-19) pandemic. Routine dental services all over the world were required to close. Those which remained open were advised to restrict treatment to urgent or emergency care only, initially through urgent dental care centres,^1^ with the overarching message being to avoid aerosol generating procedures (AGPs) wherever possible.^2-6^ Guidance on what constitutes an AGP in dentistry is not clear, and existing documents acknowledge a limited evidence base.^7^ This guidance specifically mentions high-speed dental instruments (high-speed air-turbine handpiece and ultrasonic scaler) however, orthodontic procedures are not specifically mentioned. Following the gradual reintroduction of dental services,^8-11^ significant onus is now placed on the need for appropriate personal protective equipment (PPE) wherever AGPs are carried out.^12-14^

In England, the Office of the Chief Dental Officer published its Standard Operating Procedure (SOP) for the resumption of dental services on the 4^th^ of June 2020^13^ which classified “orthodontic treatment” as a non-AGP. Guidance from a number of organisations contradicts this however, by classifying the use of a slow-speed handpiece during removal of bonded orthodontic appliances (debonding) as an AGP.^15, 16^ The British Orthodontic Society (BOS) has advised that if a clinician chooses to use a slow speed handpiece, they should work in a dry field and use high volume evacuation.^17^ The study cited by the BOS to support this recommendation showed a reduction in respirable particles of up to 44%^18^ when dental suction was used.^18^ Other authors have also demonstrated particulate aerosols produced by orthodontic debonding^19, 20^, however these studies aimed to examine dental material particulates and not droplets contaminated with saliva; additionally, these studies only focussed on the immediate vicinity of the procedure (0.1 - 0.3 m) and operator. Several methodologies have been used to evaluate dental aerosol and splatter.

These include the use of tracer dyes,^21-27^ measurement of bacterial contamination,^28-35^ and the use of optical particle counting instruments.^36, 37^ Various definitions exist for the terms “aerosol” and “splatter”. One classification described in the literature defines aerosol droplets as having a diameter of less than 10 µm,^38^ with splatter comprising droplets larger than this. Of droplets that become suspended in the air, a proportion will settle out over a varying time period (i.e. settled aerosol), and a proportion (most likely particles < 5 µm) will remain suspended (i.e. suspended aerosol). Our group has recently developed and described a reliable and valid methodology of evaluating splatter and settled aerosol created following dental procedures using a tracer dye with digital image and spectrofluorometric analysis.^39^ As part of these investigations, we have demonstrated that the use of a 3-in-1 spray (with air *and* water) produces significant contamination and should be regarded as an AGP, with a 30 second wash causing visual contamination up to one metre. The aim of the present study was to evaluate splatter and settled aerosol contamination following orthodontic debonding, including removal of composite using a slow speed handpiece with assistant-held dental suction.

## Materials and Methods

The methods used in this study have previously been described in detail elsewhere.^39^ In summary, an eight-metre diameter rig was used to support grade 1 qualitative cotton-cellulose filter papers (Whatman; Cytiva, MA, USA) spaced at 0.5 m intervals on eight, four metre rods arranged at 45-degree intervals around a dental training mannequin (Model 4820, A-dec; OR, USA). Filter papers were also placed on the forearms, chest, upper leg, and head of the operator and assistant, as well as on their masks and full-face visors. Standard hospital ventilation provided 6.5 air exchanges per hour. 2.65 mM fluorescein solution was used as a tracer and in a modification to our previous work, fluorescein was introduced into the mouth of the mannequin rather than through the water supply to model normal salivary flow. Dental models were soaked in fluorescein for 2 minutes before each experiment, and four, 10 mm diameter cotton rolls were secured to the left and right, upper and lower buccal sulci of the mannequin to replicate a natural reservoir of saliva (Figure 1). Immediately prior to starting the experiment, 5 mL of fluorescein solution was added to the labial surfaces of the teeth, and the cotton rolls. 1 mm internal diameter tubing was secured 5 mm apical to the gingival margins of the upper and lower incisors in the midline, and fluorescein was introduced through the tubing at a rate of 1.5 mL/min for the duration of the experiment to mimic the higher end of the normal stimulated salivary flow range in a healthy adult.^40^ Fluorescein was not added to the handpiece irrigation reservoir, and irrigation was not used for the procedure. Figure 1 demonstrates this setup.

**Figure 1.**
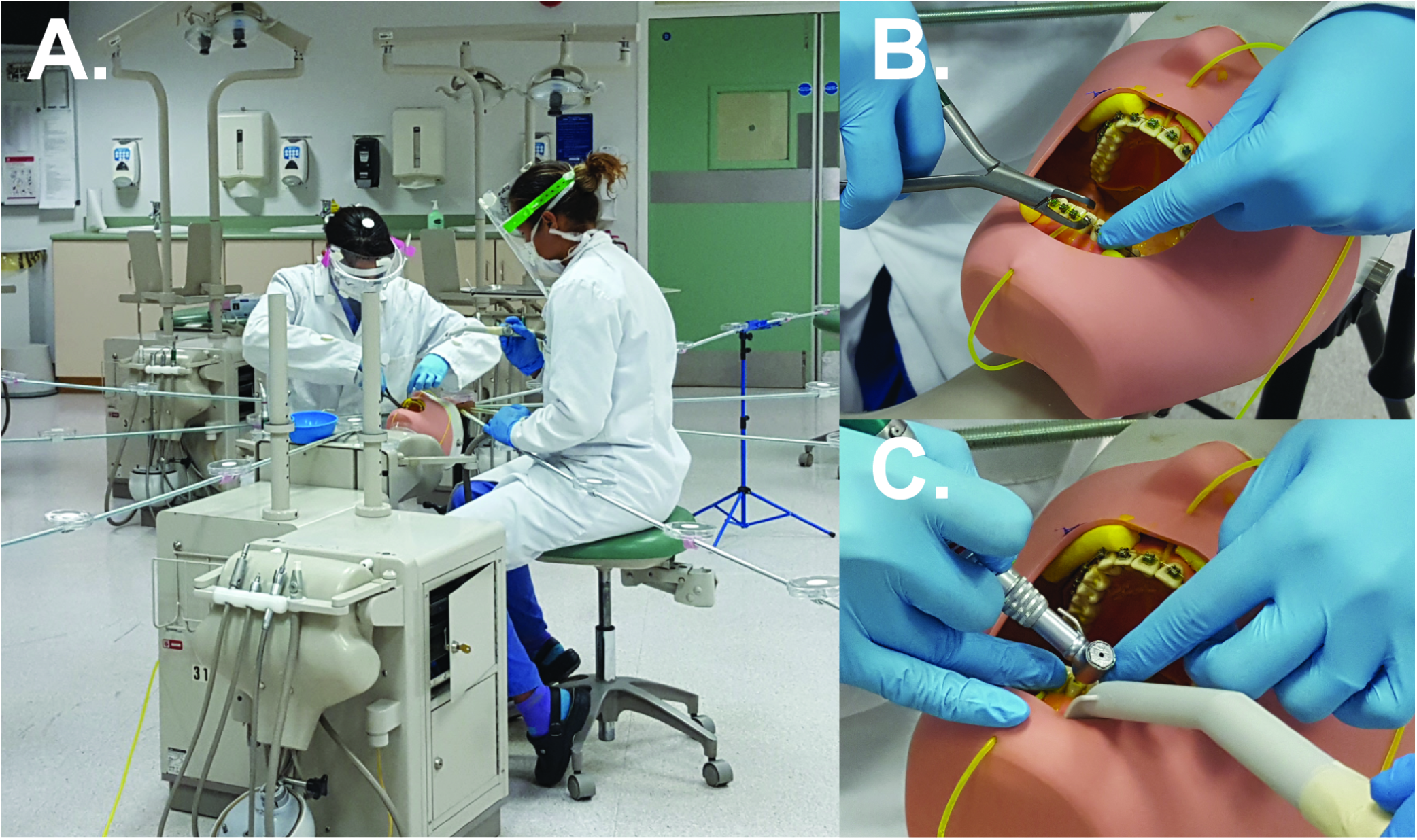
Experimental set up. A, eight-metre diameter experimental rig with operator and assistant performing orthodontic debonding procedure. B, Close up of bracket removal. Note the cotton wool rolls placed in the buccal sulci and the tubing delivering fluorescein solution. C, Close up of composite resin cement removal with slow-speed handpiece.

Prior to the experiment, GAC Ovation® (Dentsply Sirona; PA, USA) orthodontic brackets were bonded from first molar to first molar in both arches on three sets of dental models (Frasaco GmbH; Tettnang, Germany) using Unitek Transbond™ XT Light Cure Adhesive (3M UK PLC; Berkshire, UK). A macro-mechanical lock was created by preparing a 3 mm x 3 mm undercut cross in the labial surface of each tooth using an acrylic bur. This was piloted before the experiment to ensure that after removal of brackets a thin layer of composite remained on the teeth as in vivo. A 0.14 mm nickel-titanium wire was ligated in each arch. During the experiment, the brackets and archwire were removed using orthodontic debonding pliers and the teeth were polished with a fluted tungsten carbide bur in a 1:5 ratio speed-increasing dental handpiece driven by a dental air motor at full speed (with no water coolant). Large bore dental suction was operated by an assistant, closely following the operator’s handpiece, with a flow rate of 105 L/min of air measured using a commercial dental suction flow meter (RAMVAC FlowCheck, DentalEZ; PA, USA), or 6.3 L/min of water. The procedure lasted for 10 minutes and filter papers were left for 10 minutes after the end of the procedure before collection to allow settling of any splatter/ settled aerosol; this 10-minute settling time has been reported previously.^41-43^ The experiment was repeated on three separate occasions. A positive control condition – high-speed air-turbine crown preparation of the upper right central incisor with water coolant – was carried out under the same conditions as the orthodontic debonding procedure including assistant-held dental suction. This procedure was used as a positive control as it has been previously shown to produce widespread splatter and settled aerosol contamination^39^ and therefore represents a “worst case” AGP for comparison.

Contamination of the filter papers was assessed as previously described^39^ using fluorescence photography with a 500-600 nm wavelength halogen lamp (QHL75 model 503; Dentsply, NC, USA) and DSLR camera (EOS 1000D, Canon; Tokyo; Japan), and subsequent image analysis using ImageJ (v1.48 NIH; MD, USA) to give a surface area measurement (mm^2^) of fluorescein contamination; this method is likely to detect large droplets and splatter. Filter papers were also assessed using spectrofluorometric analysis as previously described^39^ by eluting fluorescein from filter papers in distilled water; a Synergy HT Microplate Reader (BioTek; VT, USA) was then used to give a quantitative fluorescence measurement in relative fluorescence units (RFU). Total RFU values for each experiment were also calculated by combining values from all samples across all replicates. This method is likely to measure the settled fraction of aerosol as well as large droplets and splatter. Examiners for both analysis methods were blinded to the experimental conditions and were calibrated for image analysis. Data were collected using Excel (2016, Microsoft; WA, USA) and analysed using SPSS (Version 24, IBM Corp.; NY, USA) using basic descriptive statistics. Heatmaps were generated using Python 3^44^.

## Results

For the positive control procedure (high-speed air-turbine crown preparation), settled aerosol and/or splatter deposition was largely concentrated in the central one metre but with some smaller deposits observed across the experimental rig (excluding mannequin, operator, and assistant). Of the 195 samples on the experimental rig (65 samples per run with three independent repeats), image analysis identified 18 contaminated samples, with spectrofluorometric analysis identifying 12 contaminated samples. There was significant operator contamination (total of 54,521 RFU across all replicates) and much less assistant contamination (total of 2,417 RFU). Combining all the samples and replicates, total contamination for the positive control was 116,482 RFU. Supplemental Table 1 provides a detailed breakdown. The pattern of contamination was similar to previously reported^39^ and is shown in Figure 2 and Supplementary Figure 1.

**Table 1.**
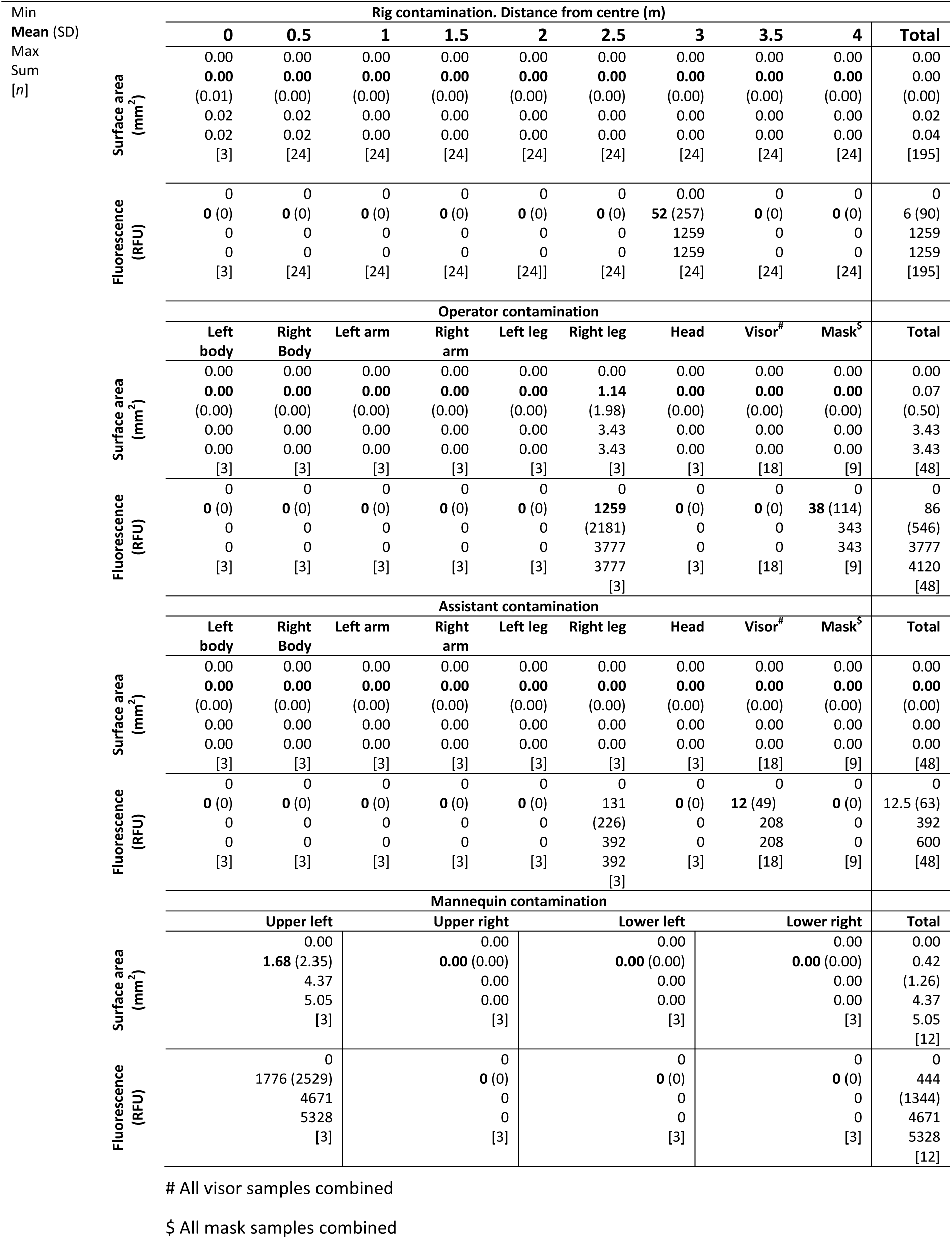
Dental aerosol and splatter as measured by contaminated surface area using image analysis or by spectrofluorometric analysis. For each experimental condition, the data from an average of three repetitions for all samples at each location are included together.

**Figure 2.**
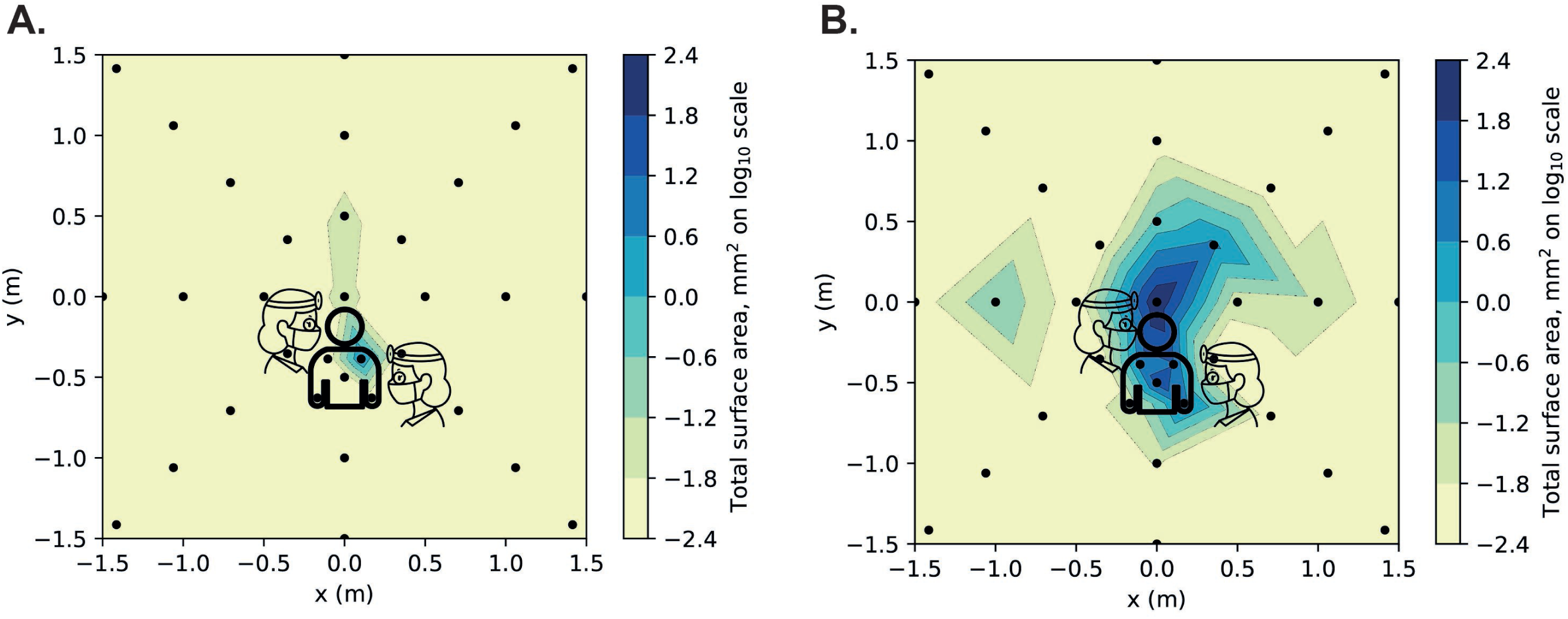
Heatmaps contaminated surface area (mm^2^) from photographic image analysis. A, Orthodontic debonding procedure. B, positive control (anterior crown preparation). For each coordinate, the maximum value recorded from three repetitions of each clinical procedure was used. Logarithmic transformation was performed on the data (Log_10_). Note the scale is reduced to remove areas showing zero readings.

For the orthodontic debonding procedure, settled aerosol and/or spatter deposition was minimal across the eight-metre diameter experimental rig. Out of 195 samples on the experimental rig, image analysis identified two contaminated samples (centre and 90° at 0.5 m). Each of these samples contained a single relevant particle, with a small surface area (0.019 mm^2^ and 0.022 mm^2^ respectively). None of these samples gave a positive reading on spectrofluorometric analysis. On spectrofluorometric analysis only one sample provided a positive result for contamination (315°, 3.5 m). See Table 1 for further details. Operator and assistant contamination was observed, with the right leg of the operator being the most contaminated location (3.43 mm^2^; 3,777 RFU) followed by the right leg of the assistant (contamination on spectrofluorometric analysis 393 RFU). The operator’s mask was contaminated on spectrofluorometric analysis only (343 RFU) as was the assistant’s visor (208 RFU). The mannequin had contamination on the upper left torso only (4.37 mm^2^; 4,671 RFU). Combining all the samples and replicates, total contamination for the orthodontic debonding procedure was 11,307 RFU.

Figures 3 and 4 present summarised data by sample location for the two procedures based on analysis technique. The orthodontic debonding procedure produced considerably less contamination across all samples. Across the experimental rig, the orthodontic debonding procedure produced 3% of the contamination of the positive control when comparing the spectrofluorometric analysis data (0% when comparing the image analysis data). Similarly, the operator had 8% of the contamination from the positive control (2% on image analysis), assistant 25% (0% on image analysis), mannequin 28% (8% on image analysis).

**Figure 3.**
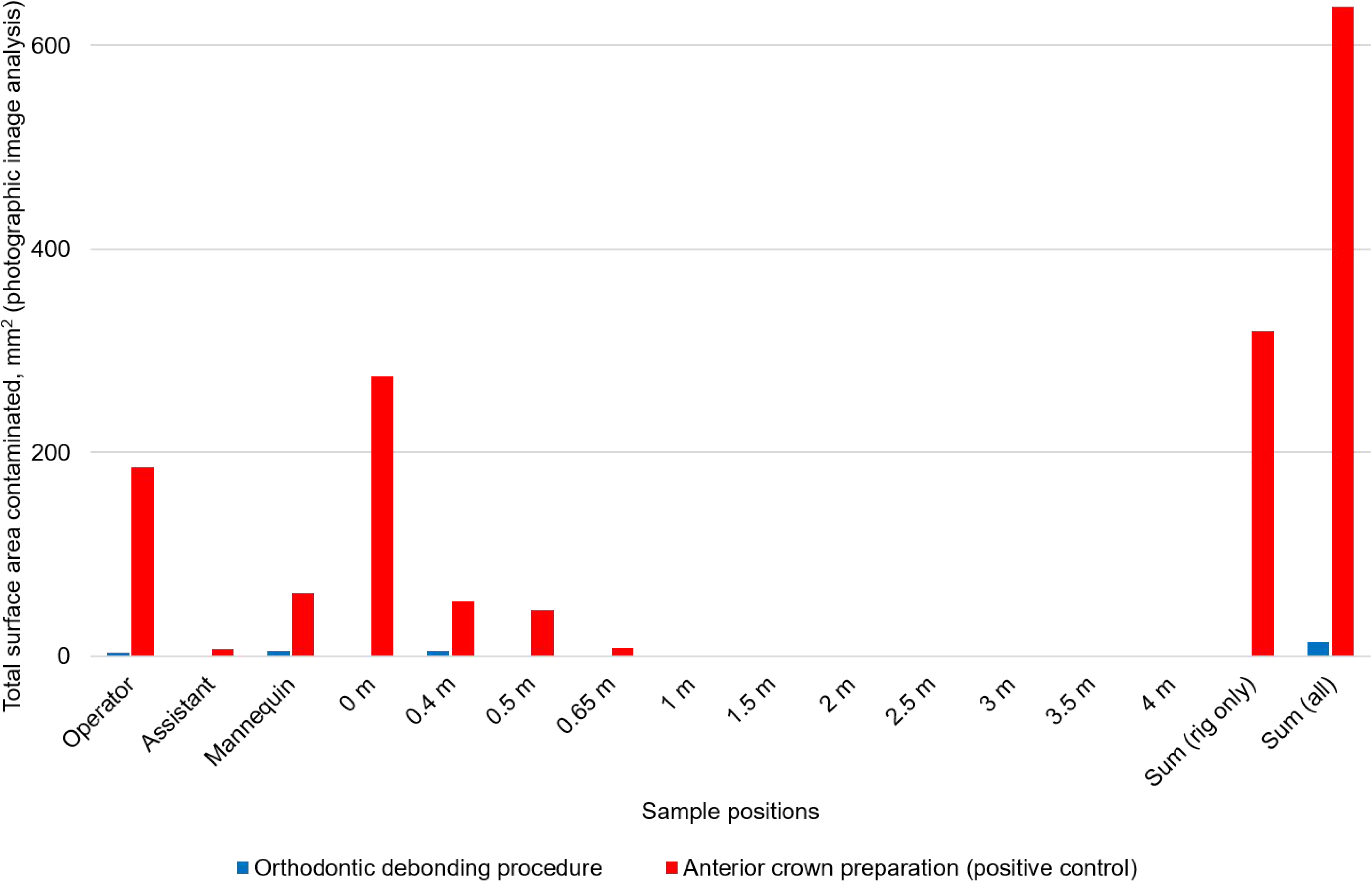
Bar chart comparing contaminated surface area (mm^2^) from photographic image analysis data by clinical procedure and sample type/location. Surface area data from each area were combined for all repetitions (i.e. all samples on the operator, assistant, mannequin, and those at 0.5m, 1m, 1.5m etc. from the centre). Note, 0.4 m and 0.65 m readings were located on the mannequin.

**Figure 4.**
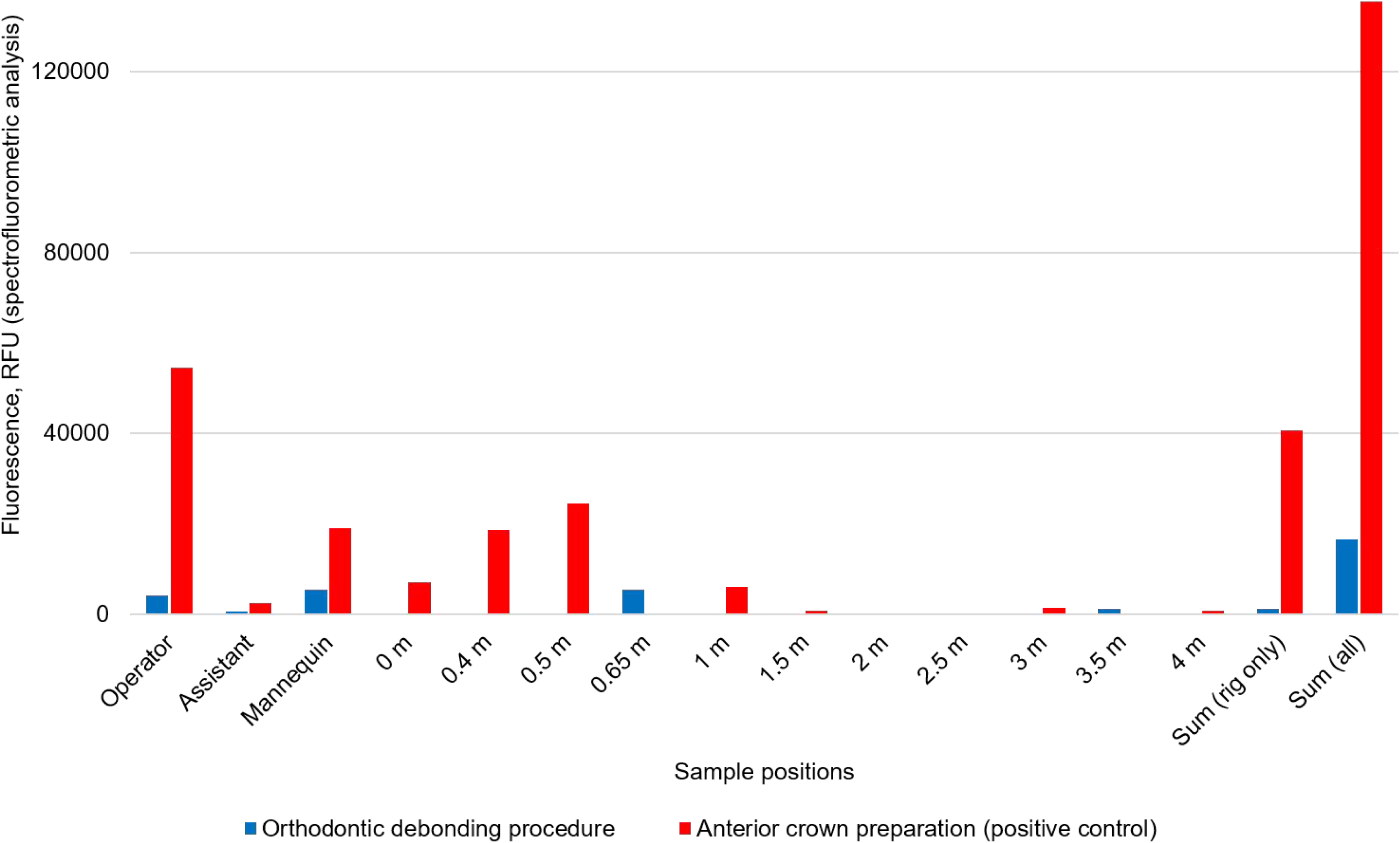
Bar chart comparing the spectrofluorometric analysis data by clinical procedure and sample type/location. RFU: relative fluorescence units. Spectrofluorometric analysis data from each area were combined for all repetitions (i.e. all samples on the operator, assistant, mannequin, and those at 0.5m, 1m, 1.5m etc. from the centre). Note, 0.4 m and 0.65 m readings were located on the mannequin.

## Discussion

The vast majority of samples showed no contamination across our eight-metre experimental area for the orthodontic debonding procedure. In contrast, our positive control (anterior crown preparation) showed much higher levels of local contamination and some distant contamination. This supports the guidance from the English CDO^13^ that an orthodontic debonding procedure using a slow-speed handpiece, in a dry field with high volume suction does not produce widespread contamination from settled aerosol and splatter. Most contamination was confined to the patient, operator, and assistant; this is consistent with microbiological studies which have investigated contamination from orthodontic debonding in the immediate vicinity of the patient, operator, and assistant.^45^

The contamination of the operator and assistant’s legs highlight the need for suitable PPE such as disposable apron, which may not have been included in PPE recommendations prior to the pandemic. The contamination on the mask, whilst probably of very low clinical significance, suggests the need to wear both a visor and a mask for adequate protection against splatter, especially if masks are used on a sessional basis. It is crucial that operating teams visually inspect each other’s masks between patients if this is the case, to ensure no visible soiling or moistening of the mask has occurred.

Our analysis identified a single positive reading (*n*= 1 of 303) at 3.5 m from the orthodontic debonding procedure, which warrants further consideration. This could be a false positive (i.e. accidental contamination), however extensive developmental work and contamination testing was completed to reduce the chance of this,^39^ and as a result we set the lower threshold of detection by spectrofluorometry at 164 RFU. The reading at 3.5 m was 1,259 RFU, eight times larger than our lower threshold (164 RFU), which suggests that this reading is unlikely to be spurious. One possible explanation for this could be related to the removal of the archwire and brackets from the mouth; a flexible wire was used which may have propelled fluorescein due to spring-back when the wire and brackets were removed from the mouth. Another possible explanation is a small volume of “saliva” (i.e. fluorescein) coming into contact with the bur. This is consistent with our previous work which demonstrated that this region (opposite the operator) is high risk for contamination across a range of dental procedures.^39^ These findings are perhaps of minor relevance for a small, closed-surgery environment but they may have wider implications for an open-plan clinic setting. Further studies should evaluate the risk of contamination in open clinical environments.

It is interesting to note that two samples (of 303) were positive for contamination on image analysis but not on spectrofluorometric analysis. One possible explanation for this is that dry debris may have settled onto these samples, but did not absorb into the filter paper, explaining the presence on photographic analysis but not on spectrofluorometric analysis (suggesting that debris may have fallen off the sample during transfer to the laboratory following photography). This is further supported by the location of these samples being very close to the source (≤ 0.5 m) in an area where dental material particulates are known to be produced from debonding.^19, 20^ Knowledge about the infectivity of severe acute respiratory syndrome coronavirus 2 (SARS-CoV-2) is still developing but from our current knowledge of the susceptibility of coronaviruses to desiccation,^46^ the risk is likely to be lower from dry debris than from droplets.

The contamination readings obtained in the present study by using fluorescein in the mouth of the mannequin were significantly lower for the positive control condition (anterior crown preparation with suction) than we have previously reported using fluorescein in the irrigation reservoirs of dental instruments.^39^ This is perhaps unsurprising but demonstrates that only a small proportion of the settled aerosol and splatter produced by dental procedures is likely to be made up of saliva (and/or blood). This dilution effect should be the subject of further study but indicates that this model is likely to be more biologically and clinically relevant.

The suction used in the present investigation would be typically described as “high flow dental suction” or “high volume evacuation”, however on the basis of the flow rate we measured, this would be more correctly classified as “medium volume suction” according to existing standards.^47^ Based on our testing (data not presented) this is within the normal range for this type of suction found in dental settings, and is consistent with previously reported studies.^34^

Our methodology has some limitations. The splatter and settled aerosol detection technique used in this study was a passive technique, in that it relied on contaminated splatter or aerosol naturally settling onto filter papers. Active sampling techniques (such as air samplers or particle counting instruments) are available and would complement this work by also allowing sampling of the proportion of the aerosol that remains suspended. However, these methods also have limitations, such as the spot nature of their sampling, which would make sampling of the 101 locations used in this investigation impractical. Also, some methods (such as optical particle meters) are non-specific, and once particle levels have returned to background levels, it is challenging to know how much of the background sample is made up of contaminated air. Because the present study relied on settling, it is likely that this methodology does not measure the fraction of aerosol which remains suspended in the air (suspended aerosol; likely droplets <5 µm). Larger droplets (up to 30 µm) may also become aerosolised and may travel some distance before settling onto surfaces;^48^ this fraction of the aerosol (settled aerosol) in addition to larger droplets and splatter will be detected by the present methodology. Both suspended and settled aerosol represent an inhalation risk, and studies addressing both components should be interpreted in combination. It is important to note, however, that emerging evidence suggests that fomite transmission may be a more significant route for SARS-CoV-2 than airborne transmission.^49, 50^ Another potential limitation of the present study is the setup of the “salivary ducts” as these were positioned in a non-anatomical position. We chose this position (Figure 1) to counteract the fact that the mannequin does not move and has no (un)conscious muscle tone or movement of the lips, tongue, and vestibules to distribute saliva around the mouth. The ducts were therefore positioned anteriorly to attempt to simulate more natural (albeit passive in our model) saliva flow around the mouth. The flow rate of instilled fluorescein (1.5 mL/min) used was also in the stimulated saliva flow range, which would likely occur when dental treatment was being performed. Our salivary flow model therefore represents a worst-case scenario for saliva contamination. Finally, as we have explored in our previous work^39^ there are still many unknowns about SARS-CoV-2 such as the infective dose. As we still do not know how viral particles are carried in aerosols produced by dental instruments and the infectivity of these, more biologically relevant models of dental bioaerosols are required.

## Conclusion

Within the limitations of this study, orthodontic debonding (with a speed-increasing handpiece, dental suction, and in settings with at least 6.5 air changes per hour) appears to produce low levels of localised settled aerosol and splatter contamination compared to a known AGP. This has implications for operator and assistant PPE (including a disposable apron, mask, and visor) and highlights the importance of patient aprons. Further work is required to examine suspended aerosol, the carriage of viral particles within dental bioaerosols, and the infectivity of these.

## Data Availability

Please contact the authors with requests for additional data.

## Acknowledgements

This study was supported by research grants from the British Endodontic Society and the Royal College of Surgeons of Edinburgh, and by the School of Dental Sciences, Newcastle University. Richard Holliday is funded by a National Institute for Health Research (NIHR) Clinical Lectureship. Charlotte Currie is funded by an NIHR Doctoral Research Fellowship. The views expressed are those of the authors and not necessarily those of the NHS, the NIHR or the Department of Health and Social Care. Nadia Rostami was funded during this period by the Dunhill Medical Trust (RPGF1810/101) who kindly extended her funding to support this urgent COVID-19 related research. We would like to thank Kimberley Pickering, Ekaterina Kozhevnikova, Jamie Coulter, and Chris Nile for their wider support on this project.

## Contributions

H. Llandro and J. R. Allison are joint first authors. H. Llandro, J. R. Allison, C. C. Currie, D. Edwards, J. Durham, N. Jakubovics, and R. Holliday, contributed to the conception and design of the study. H. Llandro, J. R. Allison, C. C. Currie, D. Edwards, C. Bowes, N. Rostami, and R. Holliday contributed to the acquisition, analysis, and interpretation of data. All authors were involved in drafting and critically revising the manuscript and have given final approval for publication. All authors agree to be accountable for all aspects of the work.

## Table/Figure Captions

**Table 1**. Settled dental aerosol and splatter as measured by contaminated surface area using image analysis or by spectrofluorometric analysis. For each experimental condition, the data from an average of three repetitions for all samples at each location are included together. RFU: relative fluorescence units.

## Supplementary Material

**Supplementary Table 1.**
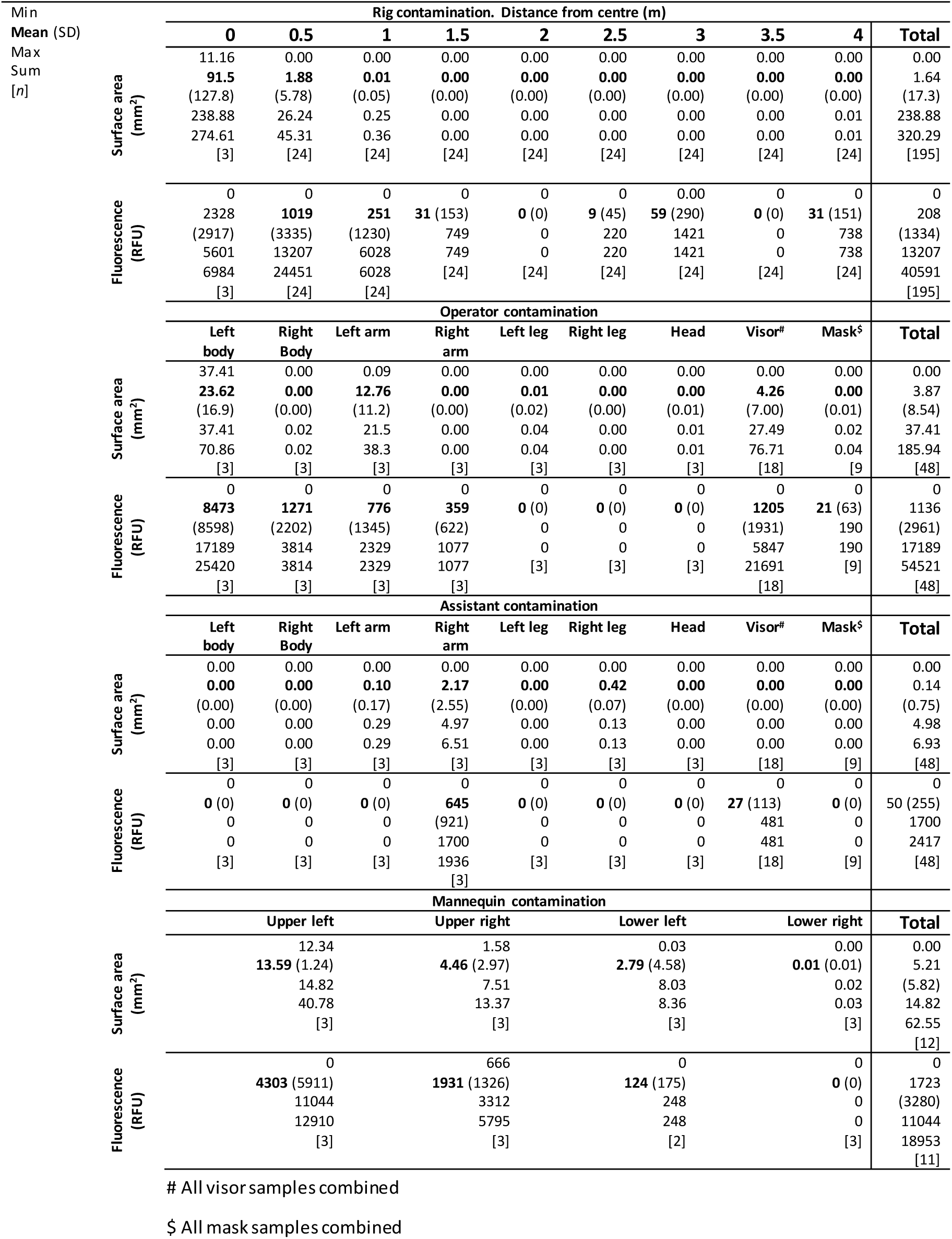
Positive control (high-speed air turbine crown preparation with assistant held dental suction). Dental aerosol and splatter as measured by contaminated surface area using image analysis or by spectrofluorometric analysis. The data from an average of three repetitions for all samples at each location are included together.

**Supplementary Figure 1.**
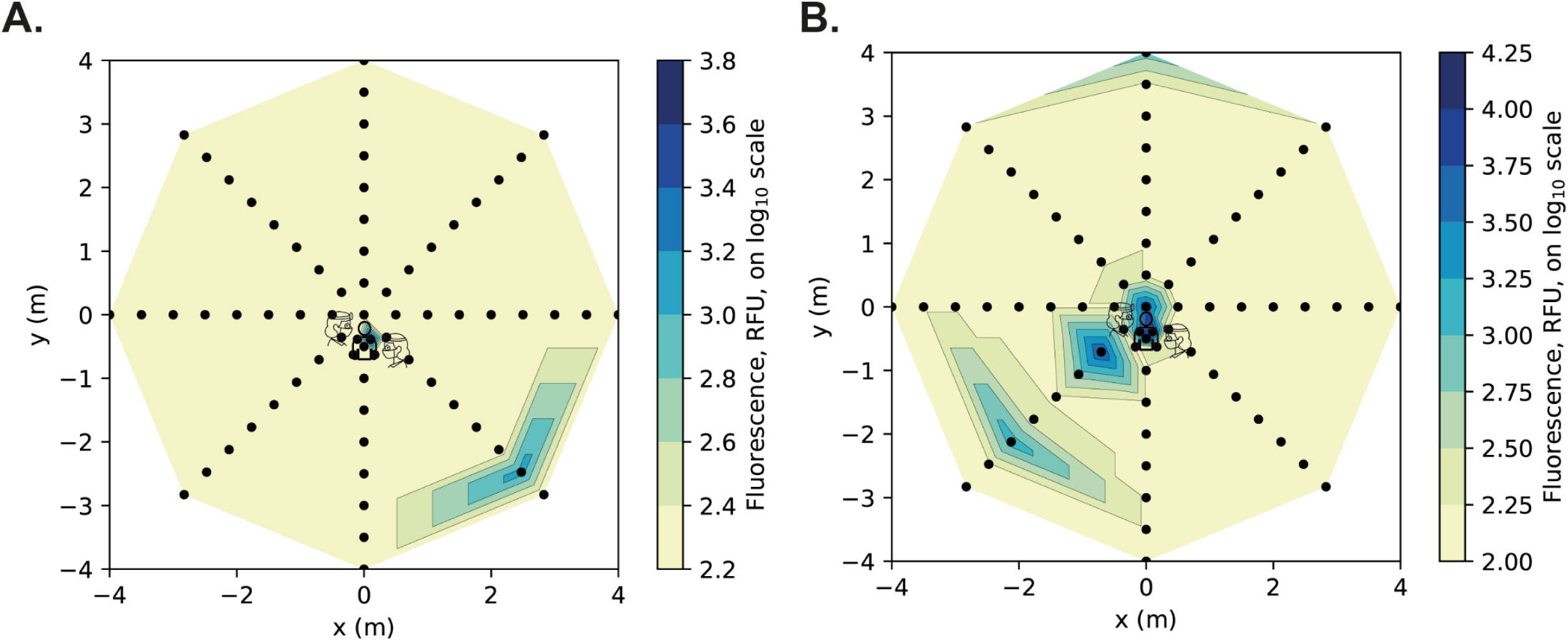
Heatmaps presenting spectrofluorometric analysis data. A, Orthodontic debonding procedure. B, positive control (anterior crown preparation). For each coordinate, the maximum value recorded from three repetitions of each clinical procedure was used. Logarithmic transformation was performed on the data (Log10). Note the scale includes the full dimensions of the experimental rig. RFU: relative fluorescence units.

